# Assessing generalizability of an AI-based visual test for cervical cancer screening

**DOI:** 10.1101/2023.09.26.23295263

**Authors:** Syed Rakin Ahmed, Didem Egemen, Brian Befano, Ana Cecilia Rodriguez, Jose Jeronimo, Kanan Desai, Carolina Teran, Karla Alfaro, Joel Fokom-Domgue, Kittipat Charoenkwan, Chemtai Mungo, Rebecca Luckett, Rakiya Saidu, Taina Raiol, Ana Ribeiro, Julia C. Gage, Silvia de Sanjose, Jayashree Kalpathy-Cramer, Mark Schiffman

**Affiliations:** Athinoula A. Martinos Center for Biomedical Imaging, Department of Radiology, Massachusetts General Hospital, Boston, MA 02129, USA; Harvard Graduate Program in Biophysics, Harvard Medical School, Harvard University, Cambridge, MA 02115, USA; Massachusetts Institute of Technology, Cambridge, MA 02139, USA; Geisel School of Medicine at Dartmouth, Dartmouth College, Hanover, NH 03755,USA; Clinical Epidemiology Unit, Clinical Genetics Branch, Division of Cancer Epidemiology and Genetics, National Cancer Institute, National Institutes of Health, Bethesda, MD 20892; Information Management Services, Calverton, MD 20705, USA; University of Washington, Seattle, WA 98195, USA; Facultad de Medicina, Universidad Mayor, Real y Pontificia de San Francisco Xavier de Chuquisaca, Sucre, Bolivia; Basic Health International, El Salvador; Cameroon Baptist Convention Health Services, Bamenda, North West Region, Cameroon; Department of Obstetrics and Gynecology, Faculty of Medicine and Biomedical Sciences, University of Yaoundé, Yaoundé, Cameroon; Department of Epidemiology, The University of Texas MD Anderson Cancer Center, Houston, TX, USA; Department of Obstetrics and Gynecology, Chiang Mai University, Chiang Mai, Thailand 50200; Department of Obstetrics and Gynecology, University of North Carolina-Chapel Hill School of Medicine, Chapel Hill, NC, USA; Beth Israel Deaconess Medical Center, Boston, United States; Department of Obstetrics and Gynaecology and South African Medical Research Council Gynaecologic Cancer Research Cancer, University of Cape Town, Cape Town; Center for Epidemiology and Health Surveillance, Oswaldo Cruz Foundation (Fiocruz), Brasília, Federal District, Brazil; MARCO Clinical and Molecular Research Center, University Hospital of Brasilia/EBSERH, Federal District, Brazil; Center for Global Health, National Cancer Institute, National Institutes of Health, Bethesda, MD 20892; ISGlobal, Barcelona, Spain; Department of Ophthalmology, University of Colorado Anschutz, Denver, Colorado, USA, 80045

**Keywords:** clinical validation, cervical cancer screening, artificial intelligence, deep learning

## Abstract

A number of challenges hinder artificial intelligence (AI) models from effective clinical translation. Foremost among these challenges are: (1) reproducibility or repeatability, which is defined as the ability of a model to make consistent predictions on repeat images from the same patient taken under identical conditions; (2) the presence of clinical uncertainty or the equivocal nature of certain pathologies, which needs to be acknowledged in order to effectively, accurately and meaningfully separate true normal from true disease cases; and (3) lack of portability or generalizability, which leads AI model performance to differ across axes of data heterogeneity. We recently investigated the development of an AI pipeline on digital images of the cervix, utilizing a multi-heterogeneous dataset (“SEED”) of 9,462 women (17,013 images) and a multi-stage model selection and optimization approach, to generate a diagnostic classifier able to classify images of the cervix into “normal”, “indeterminate” and “precancer/cancer” (denoted as “precancer+”) categories. In this work, we investigated the performance of this multiclass classifier on external data (“EXT”) not utilized in training and internal validation, to assess the portability of the classifier when moving to new settings. We assessed both the repeatability and classification performance of our classifier across the two axes of heterogeneity present in our dataset: image capture device and geography, utilizing both out-of-the-box inference and retraining with “EXT”. Our results indicate strong repeatability of our multiclass model utilizing Monte-Carlo (MC) dropout, which carries over well to “EXT” (95% limit of agreement range = 0.2 - 0.4) even in the absence of retraining, as well as strong classification performance of our model on “EXT” that is achieved with retraining (% extreme misclassifications = 4.0% for n = 26 “EXT” individuals added to “SEED” in a 2n normal : 2n indeterminate : n precancer+ ratio), and incremental improvement of performance following retraining with images from additional individuals. We additionally find that device-level heterogeneity affects our model performance more than geography-level heterogeneity. Our work supports both (1) the development of comprehensively designed AI pipelines, with design strategies incorporating multiclass ground truth and MC dropout, on multi-heterogeneous data that are specifically optimized to improve repeatability, accuracy, and risk stratification; and (2) the need for optimized retraining approaches that address data heterogeneity (e.g., when moving to a new device) to facilitate effective use of AI models in new settings.

**AUTHOR SUMMARY:** Artificial intelligence (AI) model robustness has emerged as a pressing issue, particularly in medicine, where model deployment requires rigorous standards of approval. In the context of this work, model robustness refers to both the reproducibility of model predictions across repeat images, as well as the portability of model performance to external data. Real world clinical data is often heterogeneous across multiple axes, with distribution shifts in one or more of these axes often being the norm. Current deep learning (DL) models for cervical cancer and in other domains exhibit poor repeatability and overfitting, and frequently fail when evaluated on external data. As recently as March 2023, the FDA issued a draft guidance on effective implementation of AI/DL models, proposing the need for adapting models to data distribution shifts.

To surmount known concerns, we conducted a thorough investigation of the generalizability of a deep learning model for cervical cancer screening, utilizing the distribution shifts present in our large, multi-heterogenous dataset. We highlight optimized strategies to adapt an AI-based clinical test, which in our case was a cervical cancer screening triage test, to external data from a new setting. Given the severe clinical burden of cervical cancer, and the fact that existing screening approaches, such as visual inspection with acetic acid (VIA), are unreliable, inaccurate, and invasive, there is a critical need for an automated, AI-based pipeline that can more consistently evaluate cervical lesions in a minimally invasive fashion. Our work represents one of the first efforts at generating and externally validating a cervical cancer diagnostic classifier that is reliable, consistent, accurate, and clinically translatable, in order to triage women into appropriate risk categories.

## INTRODUCTION

The development of artificial intelligence (AI) and deep learning (DL) approaches have become seemingly ubiquitous in recent years, across several clinical domains, with optimized models reporting near-clinician-level performance (1–4). However, translation of AI models from bench to bedside remain sparse. To be clinically translatable, AI/DL models should be robust, computationally-efficient, low-cost, and blend well with existing clinical workflows, ensuring the inputs/outputs of the model and the task it performs are most relevant to the clinician for a given use case. This is often not the case with existing models, which are frequently hindered by several key methodological flaws in their design (5), thereby undermining their validity, and hindering clinical translation. In particular, model robustness has emerged as a key challenge hindering AI model deployment from bench to clinical practice.

In the context of this work, model robustness refers to two key attributes: 1. repeatability or reproducibility, defined as the ability of a model to generate near-identical predictions for the same patient under identical conditions, ensuring that the model produces precise, reliable outputs in the clinical setting (6); and 2. generalizability or portability, defined as the ability of a model to adapt well to domain expansion or, alternatively, the ability of a model to perform well on datasets that are out of distribution from the training data, i.e., having different characteristics from training data (7). There is a paucity of work in the current DL and medical image classification literature that assess one or both of these attributes, with many models tending to overfit to the training data distribution. This is either due to 1) the absence of data heterogeneity (geography-, institution-, population- and/or device-level) in the available training data for a given use case; and/or 2) the absence of specific optimization approaches to reduce overfitting. To assess whether a model is overfit, an external dataset is required which has different characteristics from the training set. Assessing overfitting is particularly important when considering AI model deployment for use cases that are likely to involve multiple axes of data heterogeneity.

Globally, cervical cancer is the fourth most common cause of cancer morbidity and mortality, with approximately 90% of the 300,000 deaths per year occurring in low-resource settings (8–10). Even though the causal pathway to cervical cancer is well understood, with HPV being the main cause (11–13), this cancer has not yet been controlled, especially in low-resource settings (14). The primary prevention strategy is HPV vaccination, and for the secondary prevention strategy the World Health Organization (WHO) recommends screening with HPV test (15,16). In order to triage the risk of HPV-positive individuals, visual inspection with acetic acid (VIA) is used in low-resource settings (17,18). However, many studies have shown that expert visual evaluation has mediocre accuracy and repeatability (19,20). Therefore, there is a need for a highly accurate, repeatable, low-cost, point-of-care visual screening test to triage the risks of HPV-positive individuals. To address this need, we previously conducted a comprehensive, multi-stage model selection and optimization approach, utilizing a large, collated multi-institution, multi-device, and multi-population dataset, in order to generate a diagnostic classifier, termed automated visual evaluation (AVE) that is able to classify images of the cervix into “normal”, “indeterminate” (interchangeably termed as “gray zone”) and “precancer/cancer” (denoted as “precancer+”) categories (21). In the present work, we assess the generalizability or portability of AVE on multiple external datasets; specifically, we assess both the repeatability and classification performance, utilizing various retraining and inference strategies. Our approaches are directed by the known distribution shifts present in our external dataset, in the form of device and geography.

Our work makes two important conclusions, which, we believe, hold relevance even outside of cervical imaging:

1. Portability:

a. Device-level heterogeneity impacts model performance greater than geography level heterogeneity. Our model performs well out of the box (no retraining) on external datasets where the axis of heterogeneity is geography only vs. device, i.e., on images from a different geography but sharing a device that is represented in the training dataset.
b. Incremental retraining with inclusion of new device images to the training dataset progressively improves classification performance and class discrimination on images from a new device previously not incorporated in the training dataset, up to a point of saturation.
2. Repeatability: Our model is optimized for improved repeatability of predictions across multiple images from the same individual and continues to make repeatable predictions on external data. The repeatability performance holds true irrespective of the specific approach used for testing on the external dataset (inference vs. retraining) and the axis of data heterogeneity investigated (device vs. geography).

## MATERIALS AND METHODS

In this paper, we utilized a model that we developed in a prior study, following a multi-stage model selection and optimization process utilizing a multi-heterogeneous dataset, henceforth referred to as “SEED” (21). The primary discernible axes of heterogeneity in this prior work included image capture device and geography. In the current study, we conducted a thorough external validation of our model by running the model on images prospectively collected from a new, external dataset, henceforth termed “EXT”. The “EXT” dataset used a different image capture device, Samsung Galaxy J8, from those of the SEED (Fig. 1), and also constituted six distinct geographies/countries (Table 1, Fig. 1). All of these countries are listed in the low- and middle-income countries (LMIC) classification of the World Bank and IMF (22).

**FIGURE 1:**
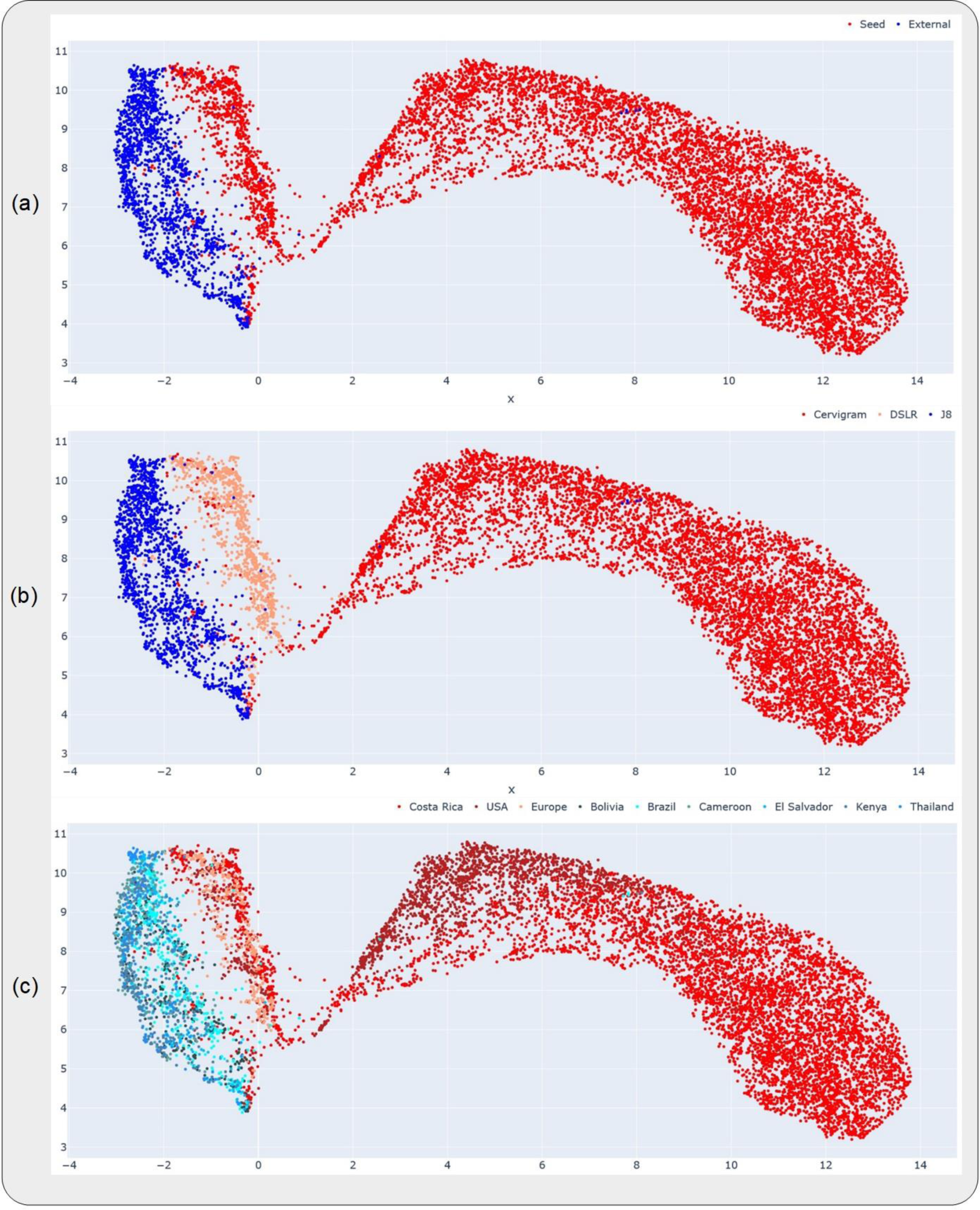
Uniform manifold approximation and projections (UMAP) highlighting the relative distributions of the datasets, devices and geographies investigated in this work. Each subplot highlights a different representation of the UMAP, where the color coding (highlighted in the corresponding legend at the top of each subplot) is at the (a) dataset-level, (b) device-level and (c) geography-level. The datasets and devices occupy distinct clusters in (a) and (b), while the geographies are all clustered together within the same device in (c).

**Table 1:**
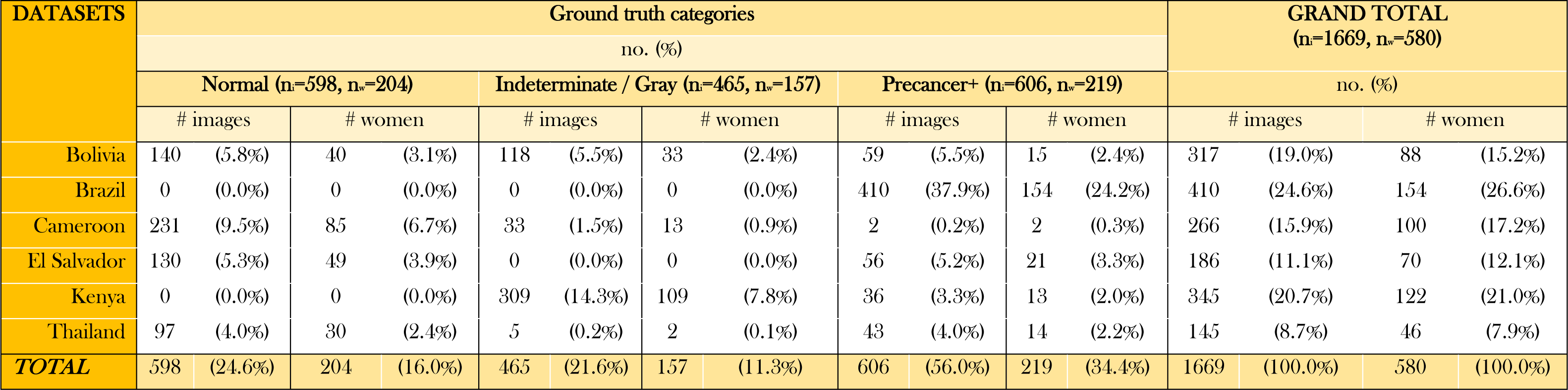
Breakdown of dataset by ground truth and geography. Detailed breakdown of “EXT” dataset by ground truth class and geography. ni=total # images; nw=total # women

### DATASET

#### Analysis Population

We utilized two groups of datasets in this study: 1) a collated, multi-institutional and multi-device (cerviscope, DSLR) dataset that was previously utilized in the model development work, which comprised of a convenience sample combining five distinct studies – Natural History Study (NHS), ASC-US/LSIL Triage Study for Cervical Cancer (ALTS), Costa Rica Vaccine Trial (CVT), Biopsy Study in the US (Biop), and Biopsy Study in Europe (D Biop) (21); we denote this dataset as “SEED”, and 2) an external multi-geography dataset of images taken by Samsung Galaxy J8 smartphones, from six countries – Bolivia, Brazil, Cameroon, El Salvador, Kenya and Thailand; we denote this dataset as “EXT”. All sites in “EXT” (except Brazil) was collected as part of the prospective AVE Network Project, where none of the images were available/used at the initial training, validation, and testing phases of the AVE algorithm. In all six countries, cervical images were collected at the vaginal exam using a Samsung Galaxy J8 smartphone. Referral for a vaginal exam was due to human papillomavirus (HPV) positivity in Bolivia and El Salvador, with additional cervical images from El Salvador collected from a randomly selected group of HPV-negative individuals. In Cameroon, Kenya, and Thailand, images were collected from VIA positive individuals at the triage visit. In Brazil, images were collected from patients with histologically confirmed cervical intraepithelial neoplasia (CIN) 2 or worse lesions. HPV tests used in these countries were Hybrid Capture 2 (HC2) (23) for Bolivia, AmpFire® (24) for Cameroon, and Care™ HPV (25) for El Salvador and Kenya. In Thailand, no HPV test was used for screening, however cytology was utilized in addition to VIA. Histopathologic confirmation of cervical cancer status in these countries were available in the form of CIN 2, CIN 3, adenocarcinoma in situ (AIS), and cervical cancer. In Brazil, images were collected after application of acetic acid and prior to Loop Electrosurgical Excision Procedure (LEEP).

#### Ground Truth Delineation

The ground truth values for the “EXT” dataset was assigned in a similar manner to that used for the “SEED” dataset (21). Specifically, the three ground truth values mapped to the images, “normal”, “indeterminate” and “precancer+”, were based primarily on histology and HPV results. All images ≥CIN 3 were assigned to precancer/cancer. If images were CIN 2, high-risk HPV (hrHPV) positivity was used to determine classes for images from all sites except Brazil: hrHPV+ was assigned to the “precancer+” class, and hrHPV-was assigned to the “indeterminate” class. All images from Brazil were >CIN 2 and were assigned to the “precancer+” class. For images where the histopathology result is <CIN 2 or missing, the ground truth class (either “normal” or “indeterminate”) was determined by a joint evaluation of a local clinician and an NCI expert colposcopist review in a site-specific manner. The final result of the ground truth distribution across each of the geographies, in terms of both the number of individuals and the number of images is depicted on Table 1.

#### Ethics

All study participants signed a written informed consent prior to enrollment and sample collection. All studies were reviewed and approved by the Institutional Review Boards of the National Cancer Institute (NCI) and the National Institutes of Health (NIH).

### MODEL TRAINING AND ANALYSIS

We conducted our model training and analysis in two distinct phases. Prior to any model runs, all images were cropped with bounding boxes generated from a YOLOv5 (26) model trained for cervix detection on the “SEED” dataset images (Fig. 2), resized to 256x256 pixels, and scaled to intensity values from 0 to 1. For the retraining runs, affine transformations were applied to the image for data augmentation.

**FIGURE 2:**
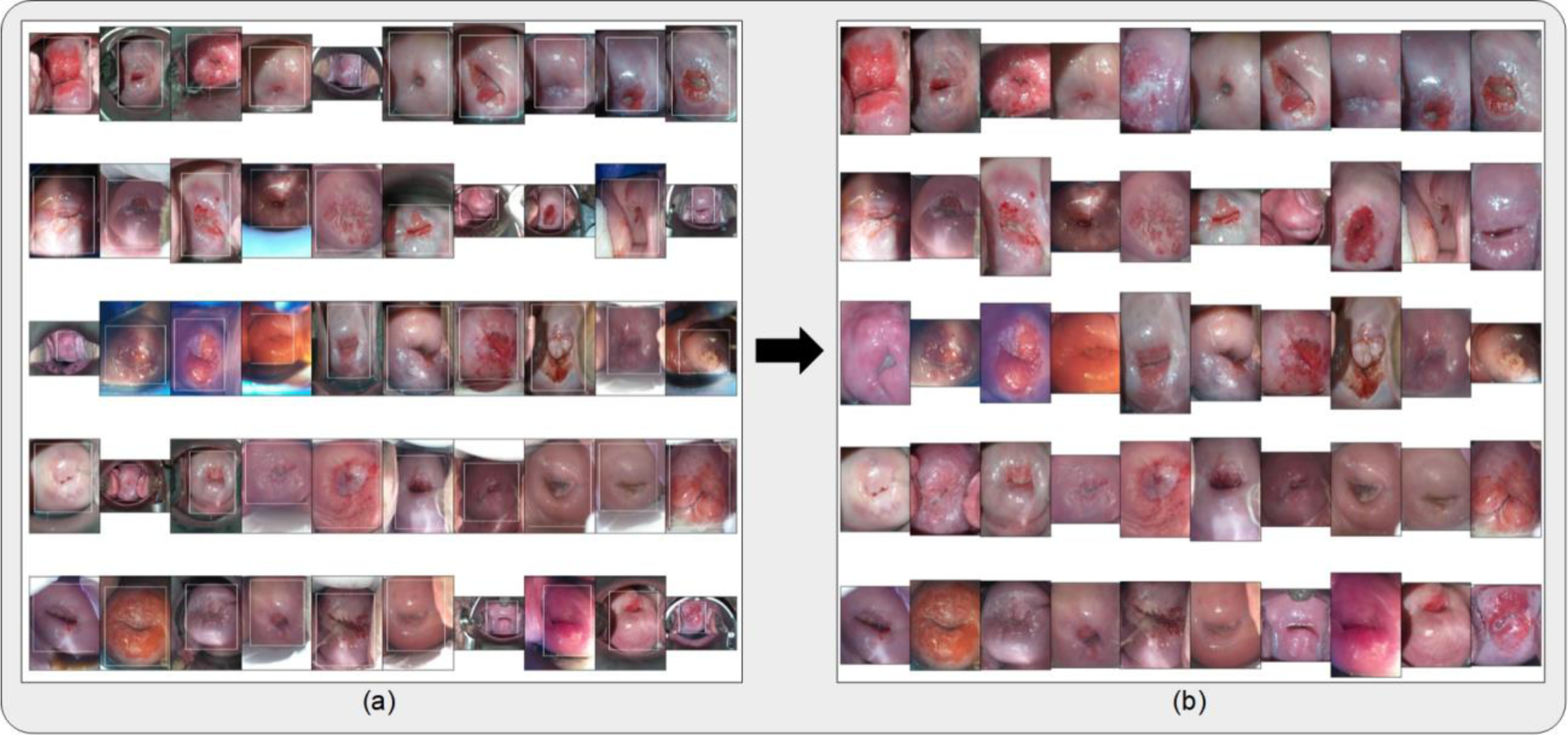
(a) Bounding boxes generated from running the cervix detector, highlighted in white, around 50 randomly selected images from the external (“EXT”) dataset. The cervix detector utilized a YOLOv5 architecture trained on the “SEED” dataset images. (b) Bound and cropped images of the cervix which are passed onto the diagnostic classifier (AVE).

#### Portability Analysis

In the first phase, we assessed the portability of our model, conducting two distinct sets of investigations. In the first set, we analyzed the relative impacts of device- and geography-level heterogeneities of our dataset on model performance, both visually via uniform manifold approximation and projection (UMAP), and statistically via assessing key classification performance and repeatability metrics.

First, in order to get a sense of the dataset distributions of the “SEED” and “EXT” datasets, including the distributions by device and geography, we ran out-of-the-box (OOB) inference with our initial model on the held-aside test set of the “SEED” dataset and on the full “EXT” dataset. We subsequently plotted UMAPs of the resulting features, which represent a dimension-reduced version of the features output from the model’s inference run, color-coded by dataset, device, and geography (Fig. 1) respectively.

We further tested the impact of device- and geography-level heterogeneity on our model performance via three distinct model runs: (i) OOB inference of AVE on a test set comprising only of “SEED” images; (ii) OOB inference of AVE on a test set comprising only of “EXT” (J8) images; and (iii) training a model using the same hyperparameters as AVE but on both “SEED” images and “EXT” images from all geographies except Bolivia and testing on Bolivia images.

In the second set of the portability analyses, we closely assessed the overall performance of AVE on “EXT” (J8) by incrementally adding women from “EXT” to our training set of “SEED” images, training on the combined set comprising of “SEED” and “EXT” images and testing on a common, held-aside set of “EXT” women (230 women, 644 images). Specifically, we added images at the woman level in two distinct ratios of ground truth – 1 normal (N) : 1 indeterminate (I) : 1 precancer+ (P), and 2 N : 2 I : 1 P; our intuition behind these additions were twofold: 1) we sought to minimize the number of precancer+ women needed when conducting a prospective study utilizing a new device, and 2) we intended to mimic the ground truth balancing utilized in our model development work, which used a 2 N : 2 I : 1 P ratio of ground truths during training on “SEED”, and evaluate whether matching the same balancing strategy as in the “SEED” set has any influence on the model performance. The specific increments of women added are highlighted in Fig. 4 and Table 2. We assessed the classification performance of the retrained models via the area under the receiver operating characteristics curve (AUROC) (Fig. 4), and the degree of extreme misclassifications (normal misclassified as precancer+ and vice versa) and total misclassifications (Table 2). We also assessed the repeatability of these models via the degree of extreme disagreement (% 2-class disagreement between image pairs across women) and the 95% limits of agreement (LoA) on a Bland-Altman plot (Table 2).

**FIGURE 3:**
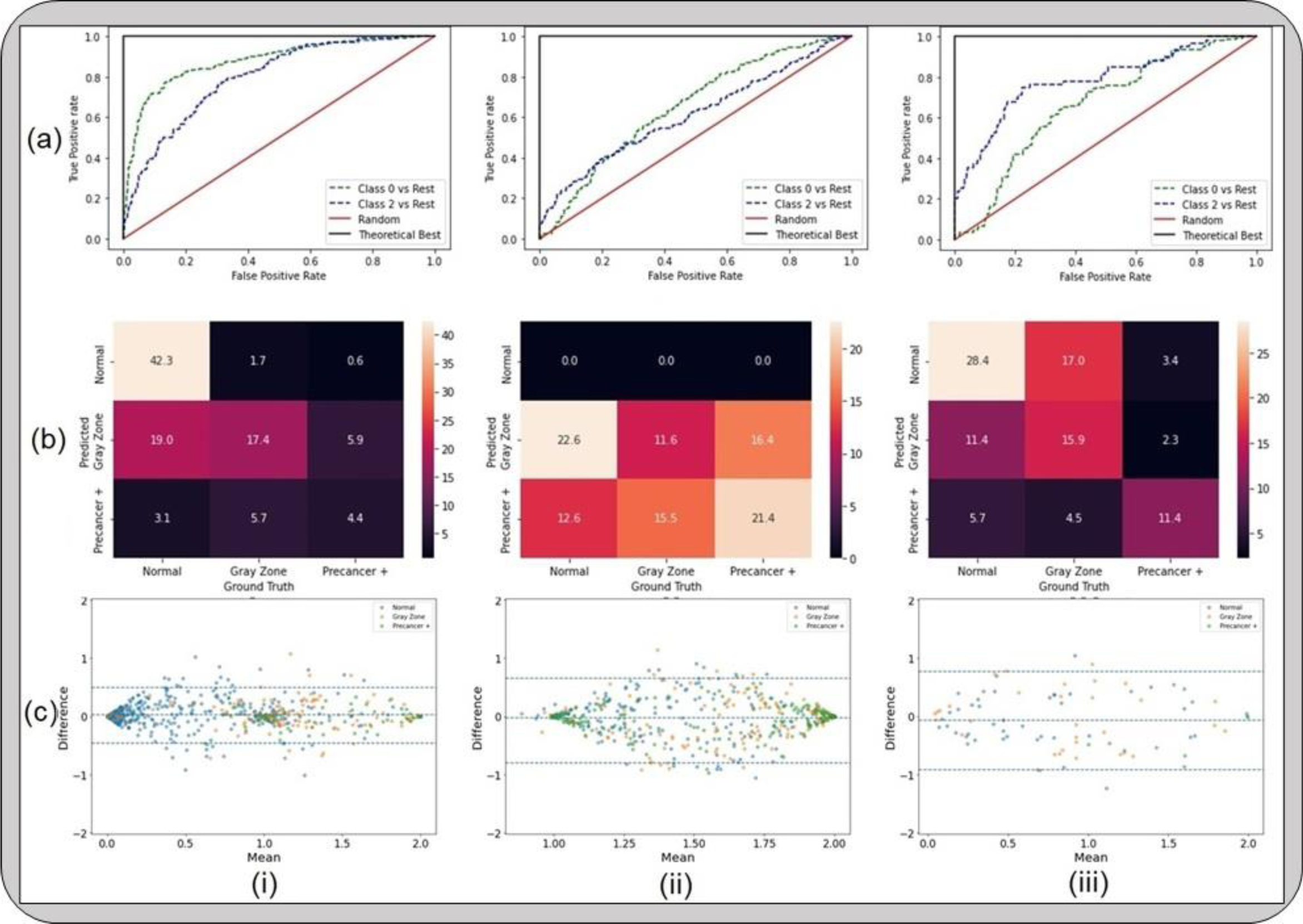
Results from the first set of the portability analyses, highlighting that device level heterogeneity impacts our model performance greater than geography level heterogeneity. The classification performance and repeatability plots depicted here include (a) receiver operating characteristics (ROC) curves; (b) confusion matrices; and (c) Bland-Altman plots, for models that were (i) trained on “SEED” and tested on a held-aside set from “SEED”; (ii) trained on “SEED” and tested on “EXT”; and (iii) trained on a dataset comprising of “SEED” + all images from “EXT” except Bolivia and tested on Bolivia images from “EXT”. “Gray Zone” = “Indeterminate” class.

**FIGURE 4:**
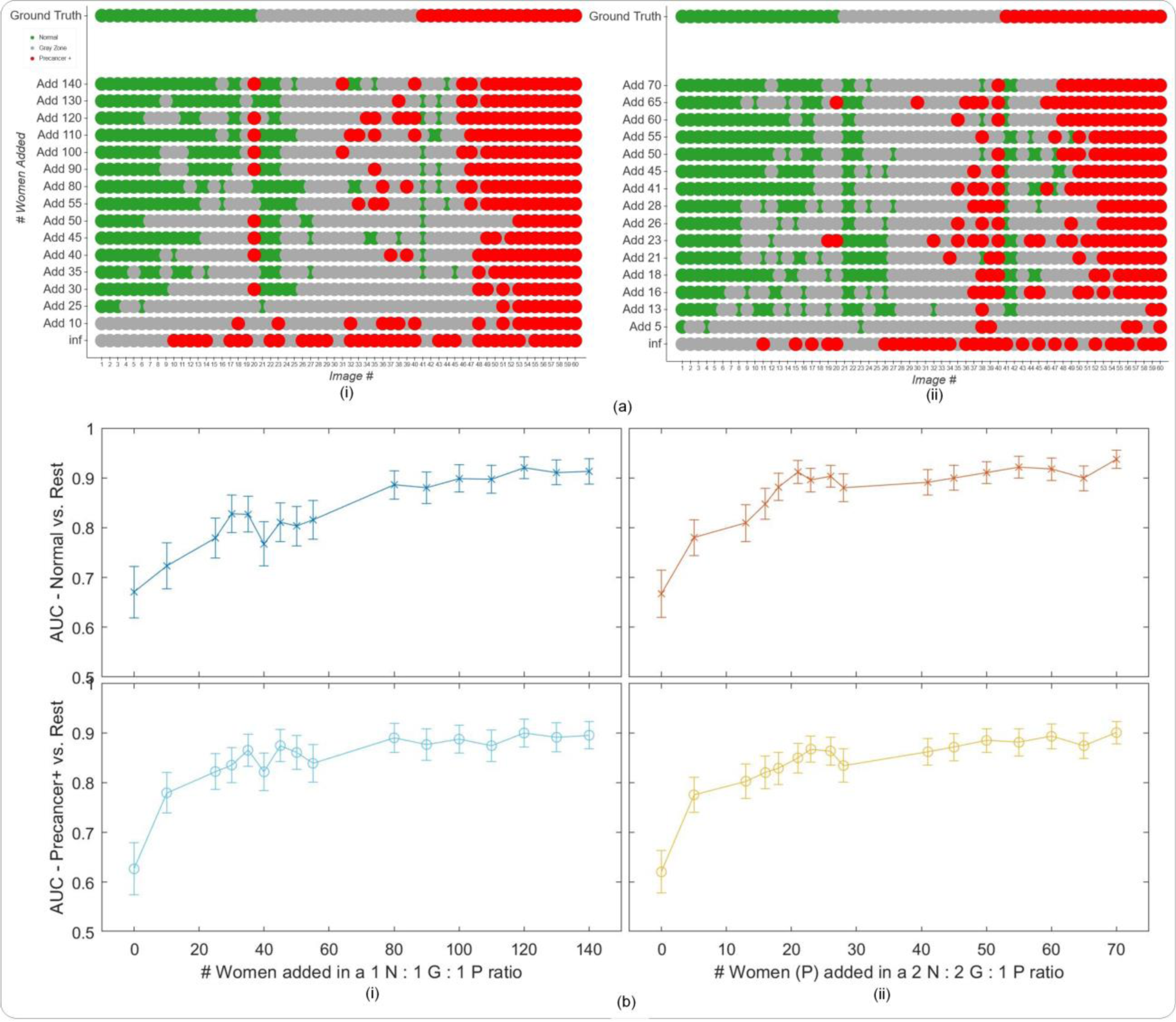
Results from the second set of the portability analysis, highlighting that retraining can improve performance on a new device previously not present in the “SEED”. (a) Model level comparison across models representing incremental additions of “EXT” (J8) images at the woman level to the training set of “SEED” images, with the “EXT” images added in (i) a 1n normal (N) : 1n indeterminate (I) : 1n precancer+ (P) ratio; and (ii) a 2n N : 2n I : 1n P ratio of ground truth classes at the woman level, where n = # of precancer+ women added (y-axes) (b) Plots of area under receiver operating characteristics curve (AUC) vs. # women added to the training set per ground truth class, in the same ratios as in (a). For example, in (ii), the x-axis represents the # precancer+ (P) women added (n) in the ratio 2n N : 2n I : 1n P to the training set. The top row plots the Normal (class 0) vs. Rest AUC, while the bottom row plots the Precancer+ (class 2) vs. rest AUC, respectively, on the y-axis. In panel (a) “normal” = green, “indeterminate” / “gray zone” = gray and “precancer+” = red.

**Table 2:** Classification and Repeatability Metrics. Relevant classification performance metrics, including % extreme misclassifications (% ext. mis.) and % total misclassifications (% tot. mis.), and repeatability metrics, including % extreme disagreement (% ext. dis.) and 95% limits of agreement (LoA) on a Bland Altman plot, for each of the model runs involving incremental additions of images from the “EXT” (J8) dataset at the woman level. Here the metrics are presented for the incremental additions in a 2n normal (N) : 2n indeterminate (I) : n precancer+ (P) ratio of ground truth class, where n = # of precancer+ women added, as shown on the leftmost column. All values are rounded to 1 decimal place.

| Table 2: Classification and Repeatability Metrics |  |  |  |  |
| --- | --- | --- | --- | --- |
| # added | Classification |  | Repeatability |  |
|  | % ext. mis. | % tot. mis. | % ext. dis. | 95% LoA |
| Add 00 (inf) |  |  |  |  |
| Add 05 |  |  |  |  |
| Add 13 | 7.8% | 65.7% | 0.0% | 0.3 |
| Add 16 | 9.6% | 53.0% | 1.9% | 0.4 |
| Add 18 | 7.9% | 55.2% | 1.0% | 0.4 |
| Add 21 | 4.4% | 51.3% | 0.5% | 0.4 |
| Add 23 | 7.4% | 39.1% | 1.0% | 0.4 |
| Add 26 | 4.8% | 46.1% | 0.0% | 0.4 |
| Add 28 | 6.0% | 55.2% | 0.5% | 0.4 |
| Add 41 | 8.7% | 37.8% | 1.9% | 0.4 |
| Add 45 | 7.0% | 44.8% | 1.5% | 0.4 |
| Add 50 | 11.3% | 39.1% | 1.9% | 0.4 |
| Add 55 | 9.6% | 38.3% | 2.4% | 0.4 |
| Add 60 | 6.5% | 33.9% | 1.9% | 0.4 |
| Add 65 | 6.0% | 39.1% | 1.0% | 0.4 |
| Add 70 | 6.0% | 27.8% | 1.9% | 0.4 |

Finally, to aid better visualization of predictions at the individual model level, we generated the plots on Fig. 4a which compare model predictions across 60 images for each of the retrained models. To generate this comparison, we first summarized each model’s output as a continuous severity *score*. Specifically, we utilized the ordinality of our problem and defined the continuous severity *score* as a weighted average using softmax probability of each class as described in Equation 3, where *k* is the number of classes and *p*_*i*_ the softmax probability of class *i*.

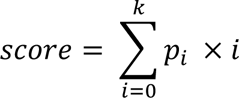

Put another way, the *score* is equivalent to the expected value of a random variable that takes values equal to the class labels, and the probabilities are the model’s softmax probability at index *i* corresponding to class label *i*. For a three-class model, the values lie in the range 0 to 2. We next computed the average of the *score* for each image across all 16 models compared and arranged the images in order of increasing *score* within each class. From this *score*-ordered list, we randomly selected 20 images per class, maintaining the distribution of mean scores within each class, and arranged the images in order of increasing average *score* within each class in the top row of Fig. 4a (i and ii), color coded by ground truth. We subsequently compared the predicted class across the models for each of these 60 images (bottom 16 rows of Figure 4a), maintaining the images in the same order as the ground truth row and color-coded by model predicted class. This enabled us to gain a deeper insight and to compare model performances at the individual image level.

#### Repeatability and Classification Performance Analysis

In the second phase of our analysis, we sought to evaluate the improvement in repeatability and classification performance imparted by the two key innovations in our AVE model, namely multiclass classification (vs. binary) and the incorporation of Monte-Carlo (MC) dropout. This was assessed by four distinct model runs which only differed in terms of these two design choices, with all other hyperparameters kept constant: (i) binary; (ii) binary with MC dropout; (iii) multiclass; and (iv) multiclass with MC dropout. Each of these models were trained on a dataset that comprised of the “SEED” dataset together with a small number of “EXT” (J8) images added in 2n N : 2n I : 1n P ratio of ground truths at the women level, for a total of n = 26 women, and tested on a held-aside “EXT” (J8) dataset. For each of these cases, we first assessed the repeatability of these models via Bland-Altman plots and the corresponding 95% LoA (Fig. 5a), test-retest score plots (Fig. 5b), and the degree of extreme disagreement (% 2-class disagreement between image pairs across women) (Fig. 5c 1) We assessed the classification performance of each of these models via the total degree of extreme misclassifications, the % precancer+ misclassified as normal, and the % normal misclassified as precancer+ (Fig. 5c 2-4).

**FIGURE 5:**
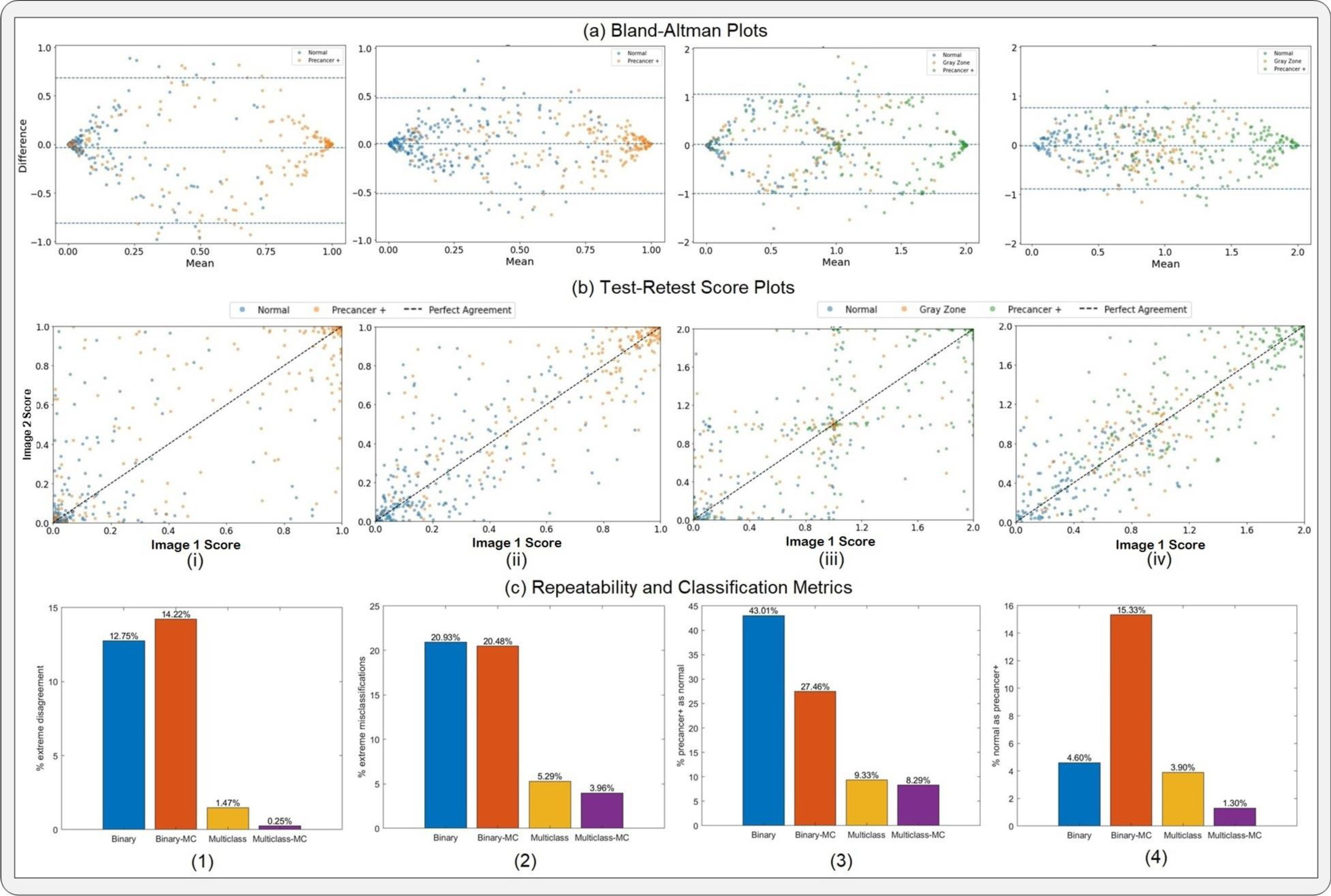
Results from the repeatability and classification performance analysis, highlighting (a) Bland-Altman plots; and (b) Test-Retest score plots for each of the four models under investigation namely (i) binary; (ii) binary with MC dropout; (iii) multiclass; and (iv) multiclass with MC dropout (our model), in order assess the relative impact of the key design choices of our model. Panel (c) (1) highlights the % extreme disagreement (proportion of women for whom the model predicts “normal” for image 1 and “precancer+” for image 2 and vice-versa) for the each of the four models (repeatability), while Panel (c) (2) – (4) highlights relevant classification metrics, including (2) the % extreme misclassification (precancer+ misclassified as normal and vice-versa); (3) the % precancer+ misclassified as normal; and (4) the % normal misclassified as precancer+, for each of the four models. “Gray Zone” = “Indeterminate” class.

## RESULTS

In this work, we conducted a multi-stage external evaluation of our model, utilizing a collated multi-geography dataset of cervical images acquired using a J8 device. We assessed portability of our model across devices and geographies, repeatability, and classification performance respectively (see METHODS).

### PORTABILITY ANALYSIS

The UMAPs on Fig. 1a and 1b highlight that the “EXT” dataset and its corresponding J8 device occupy a different cluster from the “SEED” dataset and its corresponding devices, while Fig. 1c highlights the geography level distribution. Taken together, Fig. 1a, b and c suggests the relatively greater impact of device-level heterogeneity on model performance than geography-level heterogeneity, given that within the same device, different geographies do not occupy distinct clusters on Fig. 1c, unlike the corresponding device level clusters on Fig. 1b.

This is further reinforced by Fig. 3, which highlights the results from the model runs designed to investigate the relative impacts of device- and geography-level heterogeneity. Fig. 3 illustrates that, for our model, device level heterogeneity impacts model performance greater than geography level heterogeneity. Specifically, column (i) of Fig. 3 highlights that our model performs well when running out-of-the-box inference on images that are acquired using devices that are represented in the SEED data utilized in training our model (AUROC Normal vs. Rest = 0.88, AUROC Precancer+ vs. Rest = 0.82). However, when tested on a new device, J8, out-of-the-box (OOB) inference using the same model trained on seed data fails, as indicated by the poor classification performance of our model (AUROC Normal vs. Rest = 0.65; AUROC Precancer+ vs. Rest = 0.60; no normal predictions) on column (ii) of Fig. 3. Column (iii) of Fig. 3 highlights that our model performs well when trained on images from a training set that includes the seed data and J8 images from all geographies except Bolivia and tested on J8 images from Bolivia (AUROC Normal vs. Rest = 0.70, AUROC Precancer+ vs. Rest = 0.79). This trend in classification performance is also reflected in the confusion matrices on row (b) of Fig. 3, where column (i) and column (iii) have extreme misclassification rates of 3.1% and 9.1% respectively, while column (ii) shows the model making only “indeterminate” and “precancer+” predictions, and no “normal” predictions. Finally, row (c) illustrates the strong repeatability performance of our model in all of the cases (i) – (iii), highlighted by the small width of the 95% limits of agreement (95% LoA) on each corresponding Bland-Altman plot (95% LoA = 0.24, 0.36, 0.42 respectively). Taken together, these results suggest that the classification performance of our model is affected more by device differences than differences in geography, while repeatability is relatively unaffected and quite strong throughout.

Fig. 4 illustrates that, given the impact of device level heterogeneity on the performance of our model, retraining can improve performance on a new device previously not present in the “SEED”. Specifically, incremental retraining with inclusion of J8 images to the seed data, where training set = “SEED” images + J8 images, progressively improves classification performance and class discrimination on a held-aside test set consisting only of J8 images, up until a point of saturation. Panel (a) of Fig. 4 highlights this finding by providing a detailed comparison of model performance at the individual image level. Here (i) represents the case where the J8 images were added in a 1n N : 1n G : 1n P ratio of ground truth class at the woman level, while (ii) represents J8 additions in a 2n N : 2n G : 1n P ratio of ground truth classes at the woman level (the y-axis represents n, or the number of precancer+ women added). In both cases, incremental addition of new device images to the training set improves class discrimination; this improvement is achieved with fewer precancer+ cases added to the training set in (ii), with the 2:2:1 ratio. Panel (b) plots the AUROCs (both normal vs. rest and precancer+ vs. rest) against number of women added in the training set for each of the two corresponding ratios together with bootstrapped confidence intervals for each AUROC value, further reinforcing the finding that our model performance on J8 images improves with increased representation of J8 images in the training set in a saturating fashion. Additionally, model performance on the original “SEED” set images remains consistently strong regardless of the number of women added from the “EXT” dataset (Supp. Table 1).

Table 2 highlights key classification (% extreme misclassifications and % total misclassifications) and repeatability (% extreme disagreement and 95% LoA) metrics for the case where J8 images are added to the training set in a 2 N : 2 G : 1 P ratio at the woman level. Specifically, the decrease in % total misclassifications with progressive addition of J8 images in the training set further illustrates the improvement in classification performance. On the other hand, the repeatability of our model is quite strong and relatively consistent throughout, as highlighted by the consistently low % extreme disagreement and 95% LoA values in Table 2.

### REPEATABILITY AND CLASSIFICATION PERFORMANCE ANALYSIS

We hypothesized that the two key design innovations utilized in our AVE model, namely multiclass classification (vs. binary) and the incorporation of MC dropout are optimized for both improved repeatability of predictions across multiple images from the same woman, as well as improved class discrimination, which subsequently carries over well to “external” data (“EXT”) in the form of a new device (J8). This is evidenced in Fig. 5, which highlights key classification and repeatability metrics for each of the four models under investigation namely (i) binary, (ii) binary with MC dropout, (iii) multiclass and (iv) multiclass with MC dropout (our model). Panel (a) highlights the improvement in repeatability via decrease in the 95% LoA on the Bland Altman plot; specifically, the corresponding 95% LoA values are (i) 0.75, (ii) 0.50, (iii) 0.51, and (iv) 0.41 respectively. Both the binary to multiclass transition and the no dropout to MC dropout transition improve repeatability, with the multiclass MC model achieving the best repeatability among all models. This is reinforced by the test-retest score plot in (b), which highlights progressively stronger alignment to the diagonal representing no difference between image 1 and image 2 score at the patient level, from (i) through to (iv). Panel (c) (1) also highlights that the multiclass MC model (our model) achieves the smallest degree of extreme disagreement on repeat images per woman (0.25%), i.e., the fewest women for whom image 1 is predicted “normal” and image 2 is predicted “precancer+” and vice versa. Taken together, panel (a), panel (b) and panel (c) (1) suggest that our multiclass model with MC dropout is strongly optimized for improved repeatability.

Panel (c) (2) – (4) highlight the improvement in classification performance, represented by successive decrease in % extreme misclassification, % normal misclassified as precancer+, and % precancer+ misclassified as normal, as we go from binary to binary MC to multiclass to a multiclass MC model. The incorporation of multi-level ground truth delineations during our model selection approach was designed to (1) account for the inherent clinician uncertainty or the equivocal nature of certain pathologies (e.g., ASCUS in the Bethesda system) and (2) ensure reduction of grave errors or extreme misclassifications – Panel (c) (2) – (4) highlight that this is strongly achieved, with the multiclass MC model achieving the lowest % ext. mis. = 3.96%, % p as n = 8.29%, and % n as p = 1.30% respectively (purple bars in 1 – 4).

## DISCUSSION

The use of AI models as possible biomarkers continue to be hindered by key factors that affect their clinical translation. To be effective, any biomarker needs to: 1. generate reproducible test results; 2. acknowledge uncertainty, particularly when the underlying predictive task has pre-existing uncertainty (e.g., ASCUS in the Bethesda system); and 3. acknowledge the need for, or the lack of, generalizability or portability to data heterogeneities. In this work, we address each of these properties in turn via first investigating the key axes of heterogeneities present in the underlying and prospectively acquired data, and subsequently demonstrating that the key design innovations of our multiclass AVE model are optimized for improved repeatability and classification performance and can translate well into new settings in order to facilitate clinical decision-making.

Our work demonstrates proof of principle on adapting an AI-based clinical test, which in our case was a cervical cancer screening triage test, to a new setting. Both “internal” and “external” validation of AI models, particularly for models that are intended for clinical translation and deployment across heterogeneous data, are essential for fair evaluation of model performance (27). In the context of our work, “internal” validation refers to assessing model performance on data that shares similar distributional characteristics to the training data (e.g., same device, same geography, same population), while “external” validation uses datasets that are out-of-distribution (28). In the large majority of cases, the training data that is available for an AI model is homogeneous and does not often match the intended use case. Additionally, data drift or covariate shift, a phenomenon where the distribution of input data to an AI model changes over time, can significantly impact model performance following deployment (7,29,30). This is particularly consequential in a clinical setting, where an inaccurate model prediction can lead to a cascade of potentially harmful downstream clinical decisions which might impact the health and safety of a patient. In this work, we posit that assessing AI model performance requires thorough consideration of both repeatability or reproducibility of predictions, and the discrimination ability of the model, when evaluated on “external” data from a new setting.

First, we highlight that our model achieves strong repeatability of predictions when evaluated on external data. In particular, our model makes reliable, consistent predictions on external data irrespective of the axis of data heterogeneity i.e., on individuals from new geographies or new devices. This is achieved even in the absence of retraining, and remains relatively constant throughout incremental retraining, as Table 2 highlights. This is largely attributable to the presence of MC dropout and the dedicated optimization of repeatability as a selection criterion during model optimization (6,21,31).

Second, we demonstrate that our model is able to discriminate between classes (“normal”, “indeterminate”, “precancer+”) well when evaluated on external data without retraining, provided that the axis of heterogeneity is geography only. If the external data is from a new device, our model performance improves as we incrementally add images collected from additional individuals from the external dataset and retrain on the collated training set. The specific retraining approach used, in particular, a ground truth ratio of women added to match the corresponding ratio in the “SEED” used for baseline model training, also determines the extent of this improvement. Additionally, as Fig. 3 highlights, this performance improvement eventually reaches a saturation point. Overall, these findings have important implications for clinical deployment: in order to deploy our model to a new setting which uses a different image capture device from the family of devices utilized in model training, we would need to retrain our model, via optimized strategies, with a small portion of labelled images acquired using the new device; however, this is not needed if the new setting only differs in terms of geography. We can therefore expect our model to generalize well across diverse geographies without the need for retraining, provided that the image capture device used is represented in the training set. This is a critical and impactful result, which implies that standardizing an image capture device should minimize the need for retraining.

Despite the heterogeneous nature of our dataset, our work may be limited by the number of external devices utilized. Forthcoming work will evaluate our retraining approaches and assess model performance on additional external devices. Future work will additionally optimize our model for use on edge devices, thereby promoting deployability and translation in clinical settings.

## Data Availability

The code used to train and generate results can be found at https://github.com/rknahmed0/cervix_generalizability.

## Author Contributions

Study concept and design: S.R.A., D.E., B.B., A.C.R., S.D.S., M.S., J.K.-C. Data collection: all authors. Data analysis and interpretation: all authors. Drafting of the manuscript: S.R.A, D.E. Critical revision of the manuscript for important intellectual content and final approval: all authors. Supervision: J.K.-C, M.S.

