## Supplement for "Assessing generalizability of an AI-based visual test for cervical cancer screening"

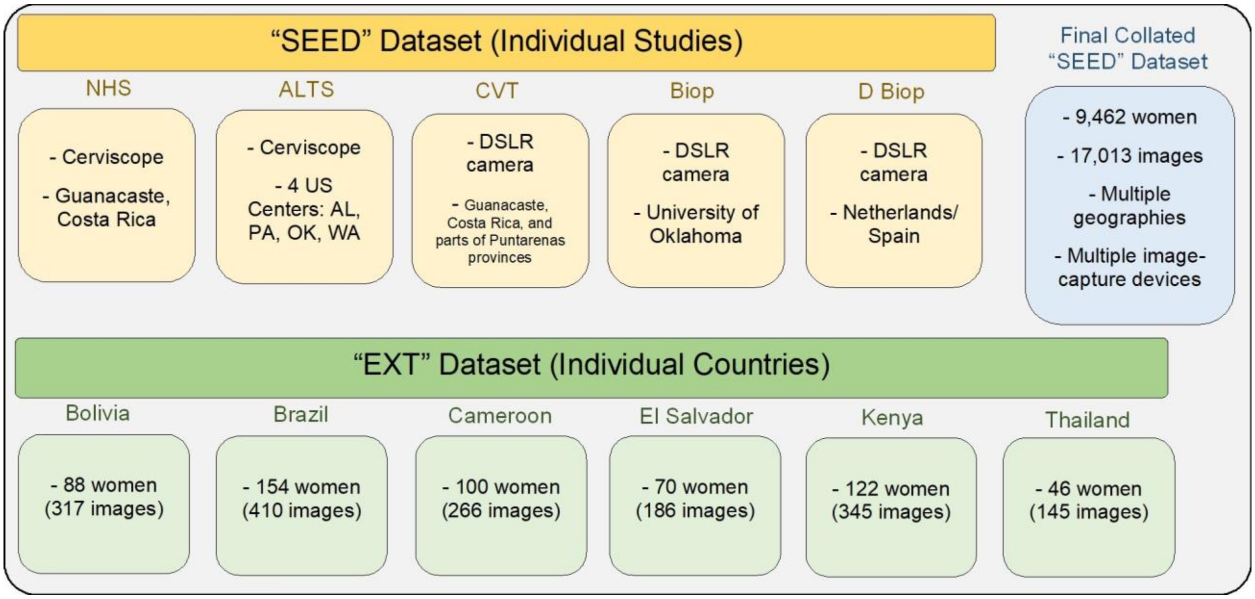


**SUPPLEMENTARY FIGURE 1:** Dataset Overview. The top panel highlights the five different studies (NHS, ALTS, CVT, Biop and D Biop) used to generate the final collated "SEED" dataset (top right) on which our model was trained and internally validated. The bottom panel highlights the six different countries / geographies included in the "EXT" dataset, all comprising of images acquired using a Samsung Galaxy J8 smartphone, on which our model was externally validated.

| Supplementary Table 1: Classification and Repeatability Metrics on "SEED" Test Set | | | | | | |
|---|---|---|---|---|---|---|
| Model # | Classification | | | | Repeatability | |
| | AUROC normal vs. rest | AUROC precancer+ vs. rest | % ext. mis. | % tot. mis. | % ext. dis. | 95% LoA |
| **Add 05** | 0.88 | 0.86 | 3.6% | 28.1% | 0.4% | 0.24 |
| Add 13 | 0.88 | 0.84 | 3.8% | 29.0% | 0.9% | 0.25 |
| Add 16 | 0.88 | 0.87 | 5.9% | 30.9% | 0.9% | 0.28 |
| Add 18 | 0.88 | 0.87 | 2.9% | 27.0% | 1.3% | 0.25 |
| Add 21 | 0.88 | 0.87 | 4.0% | 30.6% | 0.5% | 0.27 |
| Add 23 | 0.88 | 0.86 | 6.5% | 32.5% | 0.7% | 0.30 |
| Add 26 | 0.88 | 0.87 | 3.4% | 30.8% | 1.0% | 0.24 |
| Add 28 | 0.88 | 0.86 | 5.6% | 31.1% | 0.5% | 0.25 |
| Add 41 | 0.88 | 0.87 | 5.6% | 32.9% | 0.9% | 0.25 |
| Add 45 | 0.88 | 0.86 | 3.4% | 29.8% | 0.8% | 0.24 |
| Add 50 | 0.88 | 0.86 | 2.7% | 28.4% | 0.4% | 0.22 |
| Add 55 | 0.88 | 0.85 | 6.2% | 33.4% | 0.6% | 0.28 |
| Add 60 | 0.88 | 0.87 | 3.2% | 28.3% | 1.0% | 0.23 |
| Add 65 | 0.88 | 0.86 | 5.2% | 29.1% | 0.5% | 0.22 |
| Add 70 | 0.88 | 0.87 | 3.6% | 29.8% | 0.8% | 0.27 |

**SUPPLEMENTARY TABLE 1:** Classification and repeatability metrics on a held-aside test set of 8,734 images from the "SEED" dataset, highlighting that the model performs consistently well on "internal" data, even when retrained with added "external" images from the "EXT" dataset. Metrics are reported for each of the model runs involving incremental additions of images from the "EXT" (J8) dataset at the woman level, in a 2n normal (N) : 2n indeterminate (I) : 1n precancer+ (P) ratio of ground truth class, where n = # of precancer+ women added, as shown on the leftmost column. % values are rounded to 1 decimal place, while numeric values are rounded to 2 decimal places.
